# Development and external validation of machine learning algorithms for postnatal gestational age estimation using clinical data and metabolomic markers

**DOI:** 10.1101/2020.07.21.20158196

**Authors:** Steven Hawken, Robin Ducharme, Malia S.Q. Murphy, Brieanne Olibris, A. Brianne Bota, Lindsay A. Wilson, Wei Cheng, Julian Little, Beth K. Potter, Kathryn M. Denize, Monica Lamoureux, Matthew Henderson, Katelyn J. Rittenhouse, Joan T. Price, Humphrey Mwape, Bellington Vwalika, Patrick Musonda, Jesmin Pervin, AK Azad Chowdhury, Anisur Rahman, Pranesh Chakraborty, Jeffrey S.A. Stringer, Kumanan Wilson

**Affiliations:** Clinical Epidemiology Program, Ottawa Hospital Research Institute, 501 Smyth Rd, Ottawa, Canada, K1H 8L6; Clinical Epidemiology Program, Ottawa Hospital Research Institute, 1053 Carling Ave, Ottawa, Canada, K1Y 4E9; Clinical Epidemiology Program, Ottawa Hospital Research Institute, Ottawa, Canada; School of Epidemiology and Public Health, University of Ottawa, 600, Peter Morand Crescent, Ottawa, Canada, K1G 5Z3; Newborn Screening Ontario, Children’s Hospital of Eastern Ontario, 415, Smyth Rd, Ottawa, Canada, K1H 5A4; University of North Carolina at Chapel Hill, Chapel Hill, USA; University of North Carolina at Chapel Hill, Chapel Hill, USA, 27599; UNC Global Projects Zambia, Lusaka, Zambia; Department of Obstetrics and Gynaecology, University of, Zambia School of Medicine, Lusaka, Zambia; Department of Medical Statistics, University of Zambia College of, Public Health; Lusaka, Zambia; International Centre for Diarrhoeal Disease Research, 68 Shaheed, Tajuddin Ahmed Sarani, Mohakhali, Dhaka, 1212, Bangladesh; Dhaka Shishu (Children) Hospital, Sher-e-Bangla Nagar, Dhaka, 1207, Bangladesh; Department of Obstetrics and Gynecology, University of North Carolina at Chapel Hill, Chapel Hill, USA, 27599

**Keywords:** gestational age, preterm birth, metabolomics, newborn screening, prediction modelling, machine learning, validation study

## Abstract

Accurate estimates of gestational age at birth are important for preterm birth surveillance but can be challenging to reliably acquire in low and middle income countries. Our objective was to develop machine learning models to accurately estimate gestational age shortly after birth using clinical and metabolic data. We derived and internally validated three models using ELASTIC NET multivariable linear regression in heel prick blood samples and clinical data from a retrospective cohort of newborns from Ontario, Canada. We conducted external model validation in heel prick and cord blood sample data collected from prospective birth cohorts in Lusaka, Zambia (N=311) and Matlab, Bangladesh (N=1176). The best-performing model accurately estimated gestational age within about 6 days of early pregnancy ultrasound estimates in both cohorts when applied to heel prick data (MAE (95% CI) = 0.79 weeks (0.69, 0.90) for Zambia; 0.81 weeks (0.75, 0.86) for Bangladesh), and within about 7 days when applied to cord blood data (1.02 weeks (0.90, 1.15) for Zambia; 0.95 weeks (0.90, 0.99) for Bangladesh). Algorithms developed in Canada provided accurate estimates of gestational age when applied to external cohorts from Zambia and Bangladesh. Model performance was superior in heel prick data as compared to cord blood data.

## Introduction

Preterm birth is a leading cause of neonatal morbidity and mortality worldwide that disproportionately affects infants born in low- and middle-income countries (LMIC) ^1^. Knowledge of the burden of preterm birth across LMIC is essential to evaluating the impact of policies and programs aimed at improving pregnancy and neonatal outcomes. Accurate estimates of preterm birth rates rely on the use of standard definitions across jurisdictions and consistent reporting of pregnancy outcomes. International preterm birth surveillance efforts are hampered in many LMIC by limited access to ultrasound dating technology and poor recall reliability of a woman’s last menstrual period ^2–4^.The reliability of commonly used newborn assessments for estimating gestational age (GA) postnatally is often limited in preterm and growth-restricted infants, and these methods are also subject to high inter-user variability, making estimation of the burden of preterm birth in many jurisdictions problematic ^5–9^.

There is now a push by health organizations for strengthened data surveillance systems and novel tools that can more accurately assess and monitor rates of preterm birth in low resource countries, many having some of the highest preterm birth rates globally ^10,11^. Algorithms developed in North American settings by our research team and others that derive GA estimates through the biochemical analysis of newborn dried blood spots, have been shown to provide accurate estimates to within about 1-2 weeks of ultrasound-based GA ^12–15^. Our previously developed models, based on conventional multivariable linear and logistic regression methods, have been internally validated in specific ethnic subgroups of the Canadian population ^16^, and externally validated in a cohort of infants born in Matlab, Bangladesh ^17^, demonstrating satisfactory performance, but lower accuracy of GA predictions compared to the setting in which models were developed. In recent years, a number of powerful machine learning (ML) methods have been developed that allow the efficient handling of large databases, large numbers of predictors, and the incorporation of non-linear associations and complex interactions. We hypothesized that the application of advanced ML methods to postnatal metabolic GA estimation had the potential to dramatically improve model performance as compared to conventional methods. In this study, we report the performance of our newly developed ML-based GA estimation algorithms using ELASTIC NET regression ^18^ in two international prospective birth cohorts from Lusaka, Zambia, and Matlab Bangladesh.

## Methods

### Study Design and Research Participants

We sought to evaluate the performance of a GA estimation algorithm developed in a retrospective population cohort of infants from Ontario, Canada in prospective birth cohorts of infants born in Lusaka, Zambia and Matlab, Bangladesh. Detailed descriptions of all three cohorts have been published previously ^15 19 20^. A summary of the cohorts is available in Table 1.

**Table 1.** Summary of Ontario, Zambia and Bangladesh Cohorts.

| Cohort | Site | Cohort Type | Enrollment | GA Estimate |
| --- | --- | --- | --- | --- |
| **Ontario, Canada** | All hospital births and midwife attended home births | Retrospective birth registry | Infants born between January 2012 and December 2014 | First trimester ultrasound |
| **Lusaka, Zambia** | Women and Newborn Hospital of the University Teaching Hospital | Nested prospective within the Zambian Preterm Birth Prevention Study (ZAPPS) | Women <20 weeks gestation between August 2015 and September 2017 | Ultrasound at first prenatal visit |
| **Matlab, Bangladesh** | International Centre for Diarrhoeal Disease Research, Bangladesh | Nested prospective within the Preterm and Stillbirth Study, Matlab (PreSSMat) | Women 11-14 weeks gestation between January 2017 and July 2018 | Ultrasound at enrollment |

### Informed Consent and Ethical Approval

Mothers enrolled in the prospective births cohorts in Zambia and Bangladesh were approached and invited to provide informed consent to having clinical data, cord blood and newborn heel prick samples collected and sent to the Ontario Newborn Screening laboratory in Ottawa, Canada for analysis. Informed consent was provided by all participants before any data or samples were collected. Approval for the Zambia cohort was obtained from the University of Zambia School of Medicine Biomedical Research Ethics Committee (Reference number: 016-04-14), and the University of North Carolina School of Medicine (Study number: 14-2113). Approval from the Bangladesh cohort was obtained from the Research Review and Ethical Review Committees of the International Centre for Diarrhoeal Disease Research, Bangladesh (PR-16039). Approval was also obtained from the Ottawa Health Science Network Research Ethics Board (20160219-01H) and the Children’s Hospital of Eastern Ontario Research Ethics Board (16/20E) for model development in the Ontario, Canada cohort, and external validation in the Zambia and Bangladesh cohorts. All study activities were carried out in accordance with relevant institutional guidelines and regulations.

### Sample collection and analysis

Details of sample collection and analysis are included in the Online Supplement. All models were developed and internally validated based on clinical covariates and laboratory results from heel prick blood samples in the Ontario cohort. For the Zambia and Bangladesh cohorts, a subset of infants had only a heel prick or a cord blood sample collected, and a further subset had both sample types. All samples from the Ontario, Zambia and Bangladesh cohorts were analyzed centrally at the Newborn Screening Ontario (NSO) laboratory in Ottawa, Canada. Analytes were included as candidate predictors in GA estimation models based on their routine measurement as part of Ontario’s expanded newborn screening program, including hemoglobin profiles, amino acids, acylcarnitines, hormone and endocrine markers, enzymes and co-enzymes (Table 2). Management of incidental clinical findings (screen-positive cases) for conditions screened for by the NSO program identified in the course of analyzing samples have been reported elsewhere ^21,22^.

**Table 2.** Newborn screening analytes included in predictive models.

|  |  |
| --- | --- |
| **Hemoglobins** | Adult hemoglobin: HbA(A)<br>Fetal hemoglobin: HbF (F), Acetylated HbF (F1) |
| **Endocrine markers** | 17-hydroxyprogesterone (17-OHP), Thyroid stimulating hormone (TSH) |
| **Amino Acids** | Arginine (arg); phenylalanine (phe); alanine (ala); leucine (leu); ornithine (orn); citruline (cit); tyrosine (tyr); glycine (gly); methionine (met); valine (val); |
| **Acyl-carnitines** | C0; C2; C3; C4; C5; C5:1; C6; C8; C8:1; C10; C10:1; C12; C12:1; C14; C14:1; C14:2; C16; C18; C18:1; C18:2; C10:1; C12:1; C14:1; C14:2; C4OH; C5:1; C5DC; C5OH; C6DC; C16:OH; C16:1OH; C18OH; C18:1OH; C3DC; C4DC |
| **Amino Acids** | Arginine (arg); phenylalanine (phe); alanine (ala); leucine (leu); ornithine (orn); citruline (cit); tyrosine (tyr); glycine (gly); methionine (met); valine (val); |
| **Enzyme markers** | Biotinidase; immunotripsinogen |

### Statistical Analysis

All analyses were conducted using SAS 9.4 and R 3.3.2. Detailed statistical methods are provided in the Online supplement. Data preparation steps, including standardization and log transformation were applied consistently in the Ontario, Zambia and Bangladesh cohorts. A complete case analysis was used for model derivation and internal validation in Ontario due to low missingness and the large available sample size. A small proportion of subjects in the Zambia and Bangladesh cohorts had missing values for a small subset of analytes (due to sample quality degradation or insufficient blood spot volume) and these values were imputed.

Three distinct GA estimation models were derived. Models were fit and all parameters estimated using both the Ontario training subset (N=79,636) and validation subset (N=39,829) of the Ontario cohort:

**Model 1: *Baseline model*** containing only infant sex, multiple birth (yes/no), birth weight (grams), and pairwise interactions among these covariates.

**Model 2: *Analytes model*** including infant sex, multiple birth (yes/no), newborn screening analytes (listed in Table 2), and pairwise interactions among covariates.

**Model 3: *Full model*** containing infant sex, multiple birth (yes/no), birth weight (grams), newborn screening analytes, and pairwise interactions among these covariates.

Given the large number of covariates and interactions involved, Model 2 and Model 3 were fit using an ELASTIC NET ML approach. Final Ontario model equations were used to calculate an estimated GA in the test subset of the Ontario cohort that had no role in model development, as well as in the Zambia and Bangladesh external validation cohorts. For each infant, model performance was assessed by comparing the estimated GA from the model to the ultrasound-derived GA and calculating the mean absolute error (MAE) measured in weeks (the average of the absolute difference between model-based vs. ultrasound-based values across all observations). Lower MAE reflects more accurate model-based GA estimates. We also calculated the percentage of infants with GAs correctly estimated within 7 and 14 days of ultrasound-based GA. We assessed model performance overall and in important subgroups: preterm birth (<37 weeks gestation), and small-for-gestational age: below the 10^th^ (SGA10) and 3^rd^ (SGA3) percentile for birth weight within categories of gestational week at delivery and infant sex, based on INTERGROWTH-21 categories ^23^. 95% bootstrap percentile confidence intervals were calculated for all validation performance metrics, which also took account of data imputation where applicable.

### Estimated Preterm Birth Rate

We estimated the preterm birth rate by calculating the proportion of model-based GA estimates that were below 37 weeks, as well as 95% bootstrap confidence intervals, and compared these to the observed preterm birth rate in each cohort (based on ultrasound GA).

## Results

### Participant Characteristics

The Ontario internal validation (testing subset) cohort included 39,666 infants (Figure 1). In the Ontario cohort, the proportion of infants born preterm (GA<37 weeks) based on ultrasound was 5·6%, and 3·9% and 0·92% of infants were classified as SGA10 and SGA3, respectively. In the external validation cohorts, a total of 142 heel prick samples and 265 cord blood samples collected from 311 unique newborns from Zambia, and 520 heel prick samples and 1139 cord blood samples collected from 1176 unique newborns from Bangladesh. 32 (22·5%) heel and 99 (37·4%) cord samples from Zambia were missing one or more analytes, with a maximum of three missing in any sample. 28 (5·4%) heel and 22 (1·9%) cord samples from Bangladesh were missing one or more analytes, with a maximum of 5 and 3 missing values respectively (one subject missing all values was removed from the analysis). Based on ultrasound GA, the preterm birth proportions were similar in the Zambia and Bangladesh cohorts (9·6% versus 9·7 %). The Zambia cohort had a much lower proportion of newborns classified as SGA10 as compared to the Bangladesh cohort (9·7% vs. 25·3%). Characteristics of unique infants in each cohort, as well as the infants represented in heel prick and cord blood sample cohorts are provided in Table 3.

**Table 3.**
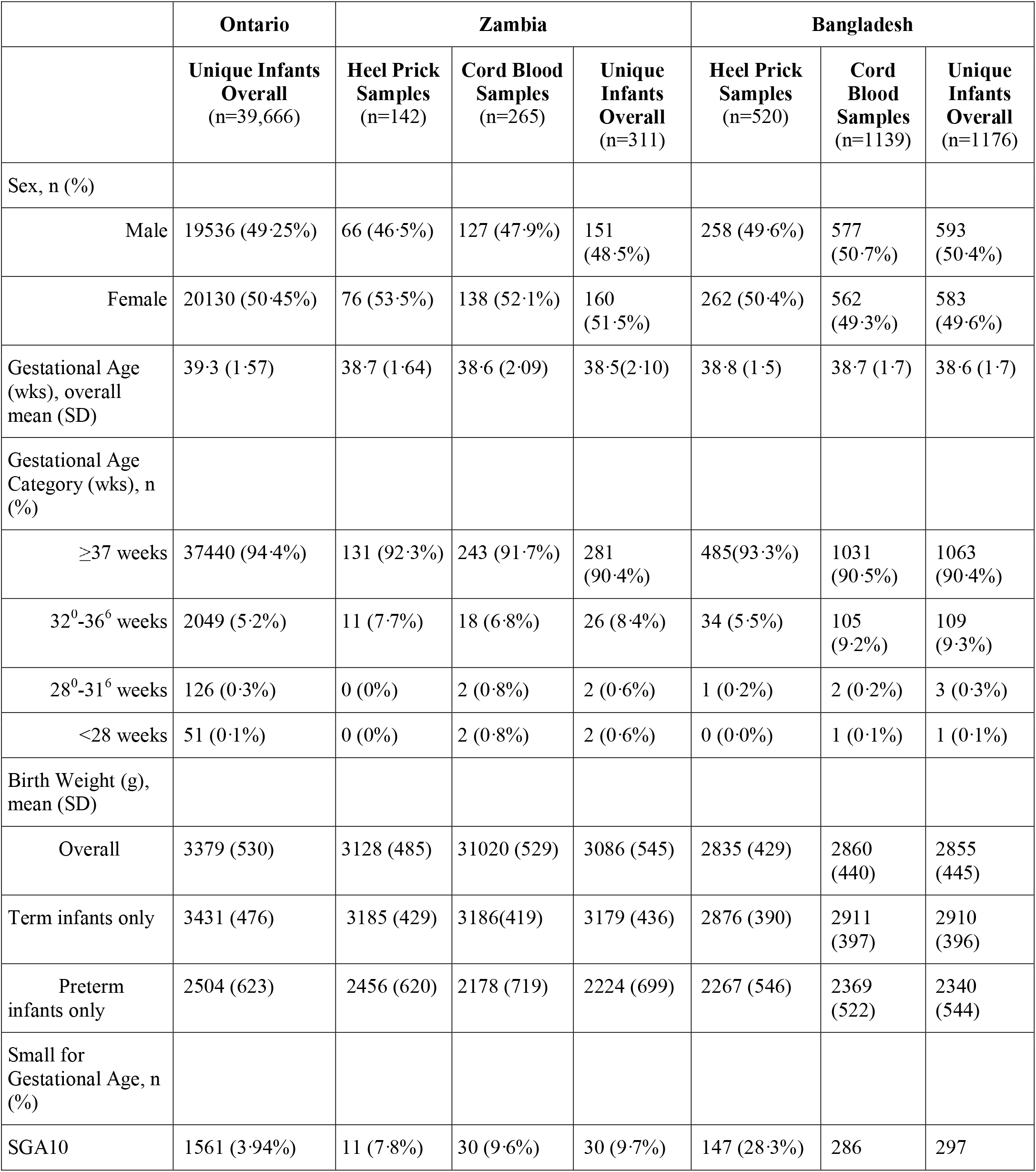
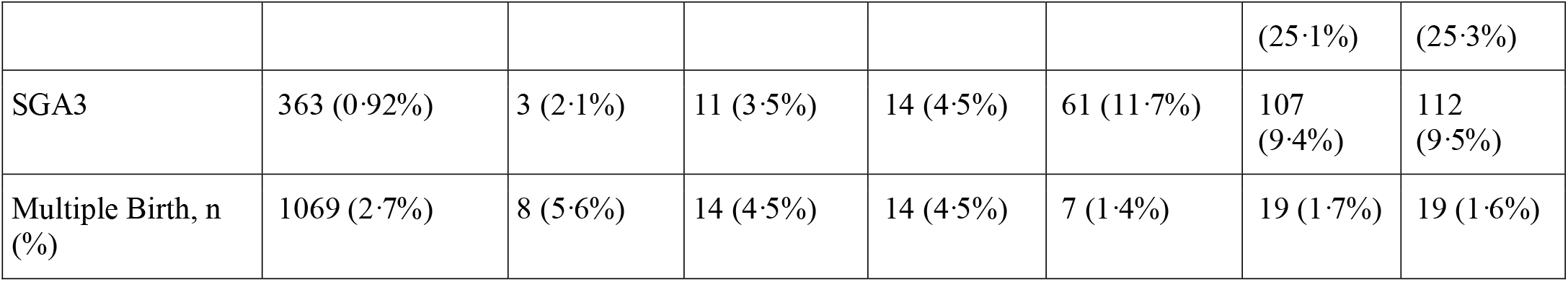
Infant Characteristics.

**Figure 1:**
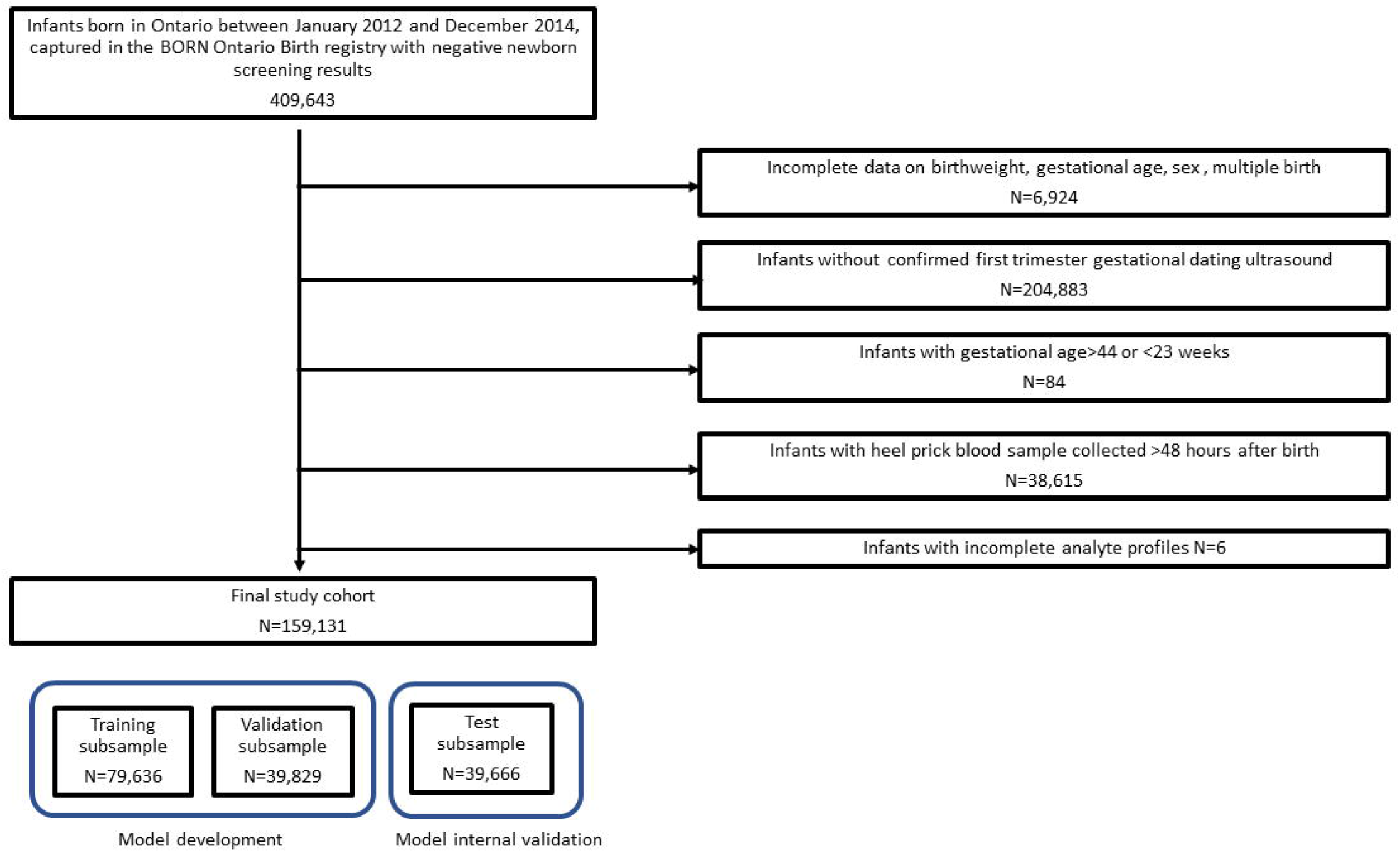
Cohort creation.

### Model performance using heel prick data from Ontario, Zambia and Bangladesh

Model external validation performance was largely similar between the Zambia and Bangladesh cohorts. Accuracy of estimated GA was generally lower in the external validation cohorts than in the Ontario internal validation results for the same models. In general, Model 3, which included both clinical and analyte predictors, outperformed Models 1 and 2. Accuracy of GA estimates in all models was highest in term infants and tended to be lower in preterm and SGA infants, but Model 3 consistently provided the most accurate estimates across the spectrum of GA (Table 4 and Figure 2). Model 1 estimated GA to within approximately 7 days of ultrasound-assigned GA, with similar performance among samples from Ontario, Zambia and Bangladesh (MAE (95% CI) 0·96 (0·95,0·97), 0·96 (0·82,1·12) and 0·99 (0·92,1·06) weeks, respectively). Model 3 estimated GA to within 5 days of ultrasound-assigned values when applied to the Ontario data (MAE (95% CI) 0·71 (0·71, 0·72) and within 6 days in the Zambia and Bangladesh data (MAE (95% CI) 0·79 (0·69, 0·90) and 0·81 (0·75, 0·86), respectively). GA was correctly estimated to within 1 week of ultrasound-assigned values for 74·6% (95% CI 74·2,75·1), 69·4% (60·6, 77·5) and 68·4% (64·3, 72·4) of heel prick samples from Ontario, Zambia and Bangladesh, respectively, and within two weeks for over 95% of subjects across all 3 cohorts.

**Table 4.**
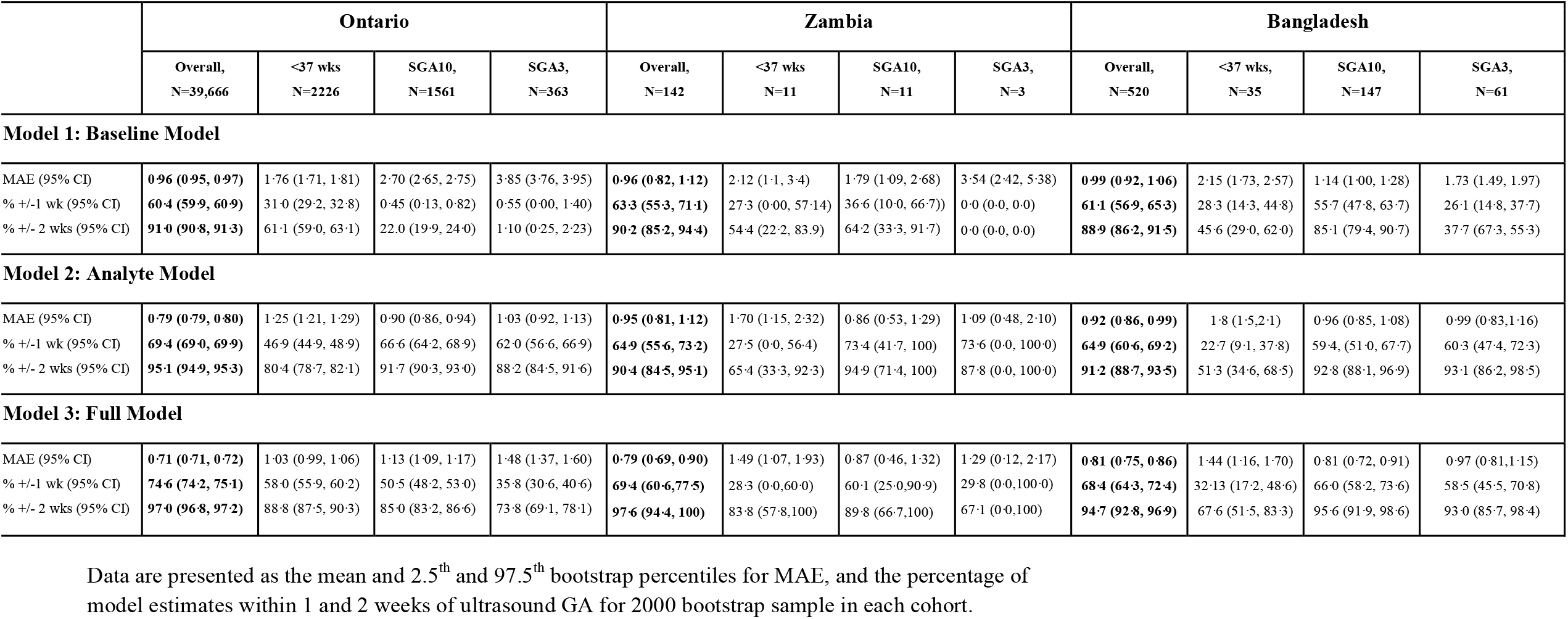
Heel prick samples: MAE and proportion estimated within 1 week, 2 weeks of ultrasound-assigned gestational age.

|  | Ontario |  |  |  | Zambia |  |  |  | Bangladesh |  |  |  |
| --- | --- | --- | --- | --- | --- | --- | --- | --- | --- | --- | --- | --- |
|  | Overall,<br>N=39,666 | <37 wks<br>N=2226 | SGA10,<br>N=1561 | SGA3,<br>N=363 | Overall,<br>N=142 | <37 wks<br>N=11 | SGA10,<br>N=11 | SGA3,<br>N=3 | Overall,<br>N=520 | <37 wks,<br>N=35 | SGA10,<br>N=147 | SGA3,<br>N=61 |
| **Model 1: Baseline Model** |  |  |  |  |  |  |  |  |  |  |  |  |
| MAE (95% CI) | **0·96 (0·95, 0·97)** | 1·76 (1·71, 1·81) | 2·70 (2·65, 2·75) | 3·85 (3·76, 3·95) | **0·96 (0·82, 1·12)** | 2·12 (1·1, 3·4) | 1·79 (1·09, 2·68) | 3·54 (2·42, 5·38) | **0·99 (0·92, 1·06)** | 2·15 (1·73, 2·57) | 1·14 (1·00, 1·28) | 1·73 (1·49, 1·97) |
| % +/-1 wk (95% CI) | **60·4 (59·9, 60·9)** | 31·0 (29·2, 32·8) | 0·45 (0·13, 0·82) | 0·55 (0·00, 1·40) | **63·3 (55·3, 71·1)** | 27·3 (0·00, 57·14) | 36·6 (10·0, 66·7)) | 0·0 (0·0, 0·0) | **61·1 (56·9, 65·3)** | 28·3 (14·3, 44·8) | 55·7 (47·8, 63·7) | 26·1 (14·8, 37·7) |
| % +/- 2 wks (95% CI) | **91·0 (90·8, 91·3)** | 61·1 (59·0, 63·1) | 22·0 (19·9, 24·0) | 1·10 (0·25, 2·23) | **90·2 (85·2, 94·4)** | 54·4 (22·2, 83·9) | 64·2 (33·3, 91·7) | 0·0 (0·0, 0·0) | **88·9 (86·2, 91·5)** | 45·6 (29·0, 62·0) | 85·1 (79·4, 90·7) | 37·7 (67·3, 55·3) |
| **Model 2: Analyte Model** |  |  |  |  |  |  |  |  |  |  |  |  |
| MAE (95% CI) | **0·79 (0·79, 0·80)** | 1·25 (1·21, 1·29) | 0·90 (0·86, 0·94) | 1·03 (0·92, 1·13) | **0·95 (0·81, 1·12)** | 1·70 (1·15, 2·32) | 0·86 (0·53, 1·29) | 1·09 (0·48, 2·10) | **0·92 (0·86, 0·99)** | 1·8 (1·5,2·1) | 0·96 (0·85, 1·08) | 0·99 (0·83,1·16) |
| % +/-1 wk (95% CI) | **69·4 (69·0, 69·9)** | 46·9 (44·9, 48·9) | 66·6 (64·2, 68·9) | 62·0 (56·6, 66·9) | **64·9 (55·6, 73·2)** | 27·5 (0·0, 56·4) | 73·4 (41·7, 100) | 73·6 (0·0, 100·0) | **64·9 (60·6, 69·2)** | 22·7 (9·1, 37·8) | 59·4, (51·0, 67·7) | 60·3 (47·4, 72·3) |
| % +/- 2 wks (95% CI) | **95·1 (94·9, 95·3)** | 80·4 (78·7, 82·1) | 91·7 (90·3, 93·0) | 88·2 (84·5, 91·6) | **90·4 (84·5, 95·1)** | 65·4 (33·3, 92·3) | 94·9 (71·4, 100) | 87·8 (0·0, 100·0) | **91·2 (88·7, 93·5)** | 51·3 (34·6, 68·5) | 92·8 (88·1, 96·9) | 93·1 (86·2, 98·5) |
| **Model 3: Full Model** |  |  |  |  |  |  |  |  |  |  |  |  |
| MAE (95% CI) | **0·71 (0·71, 0·72)** | 1·03 (0·99, 1·06) | 1·13 (1·09, 1·17) | 1·48 (1·37, 1·60) | **0·79 (0·69, 0·90)** | 1·49 (1·07, 1·93) | 0·87 (0·46, 1·32) | 1·29 (0·12, 2·17) | **0·81 (0·75, 0·86)** | 1·44 (1·16, 1·70) | 0·81 (0·72, 0·91) | 0·97 (0·81,1·15) |
| % +/-1 wk (95% CI) | **74·6 (74·2, 75·1)** | 58·0 (55·9, 60·2) | 50·5 (48·2, 53·0) | 35·8 (30·6, 40·6) | **69·4 (60·6,77·5)** | 28·3 (0·0,60·0) | 60·1 (25·0,90·9) | 29·8 (0·0,100·0) | **68·4 (64·3, 72·4)** | 32·13 (17·2, 48·6) | 66·0 (58·2, 73·6) | 58·5 (45·5, 70·8) |
| % +/- 2 wks (95% CI) | **97·0 (96·8, 97·2)** | 88·8 (87·5, 90·3) | 85·0 (83·2, 86·6) | 73·8 (69·1, 78·1) | **97·6 (94·4, 100)** | 83·8 (57·8,100) | 89·8 (66·7,100) | 67·1 (0·0,100) | **94·7 (92·8, 96·9)** | 67·6 (51·5, 83·3) | 95·6 (91·9, 98·6) | 93·0 (85·7, 98·4) |

Data are presented as the mean and 2.5<sup>th</sup> and 97.5<sup>th</sup> bootstrap percentiles for MAE, and the percentage of model estimates within 1 and 2 weeks of ultrasound GA for 2000 bootstrap sample in each cohort.

**Figure 2:**
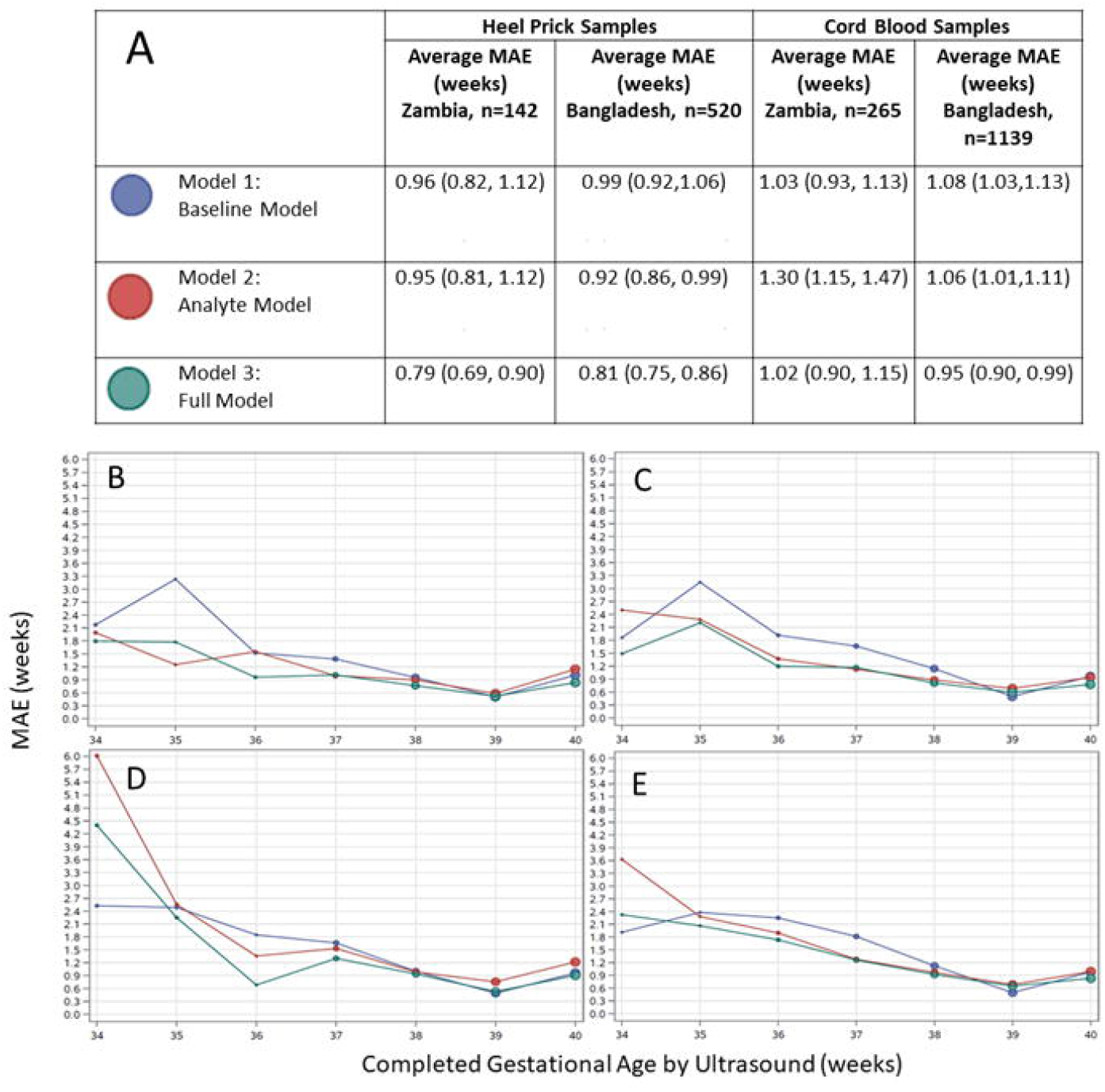
Agreement between algorithmic gestational age estimations compared to ultrasound-assigned gestational age. (A) Legend, and overall MAE (95% CI) for heel prick sample and cord blood samples from Zambia and Bangladesh across three models. Dot size in plots is proportional to sample size in each gestational age category. Performance of each model by ultrasound-validated gestational age when applied to heel prick data in (B) Zambia (C) Bangladesh, and cord blood data in (D) Zambia (E) Bangladesh. MAE, mean absolute error (average absolute deviation of observed vs. predicted gestational age in weeks).

When applied to samples from preterm infants in Ontario, Model 3 correctly estimated GA to within 7 days of ultrasound-assigned values (MAE (95%CI) 1·03 (0·99, 1·06)), and performed significantly better than Models 1 and 2 (MAE (95%CI) 1·76 (1·71, 1·81) and 1·25 (1·21, 1·29), respectively). The number of preterm infants in the external validation cohorts was small, with only 11 heel prick samples from Zambia and 35 from Bangladesh, however in both settings, model 3 outperformed both models 1 and 2, and was accurate to within about 10 days on average in preterm infants (MAE (95% CI) 1·49 (1·07, 1·93) and 1·44 (1·16, 1·70), respectively in Zambia and Bangladesh).

When applied to Ontario heel prick samples from SGA infants, Model 2 had the best performance, estimating GA to within about 6 days of ultrasound-assigned values for SGA10 and 7 days for SGA3 subgroups (MAE (95% CI) 0·90 (0·86, 0·94) and 1·03 (0·92, 1·13), respectively). Our validation cohort from Zambia contained only 11 SGA10 and 3 SGA3 classified infants. When applied to these samples, Models 2 and 3 both outperformed Model 1, and estimated GA to within about 6 days of ultrasound-assigned values in SGA10 infants, and within about 9 days in SGA3 infants. Our heel prick sample cohort from Bangladesh contained a larger number of SGA samples (147 SGA10 and 61 SGA3 samples). When applied to these samples from Bangladesh, Model 3 estimated GA to within about 6 days of ultrasound-assigned values in the SGA10 subgroup and within about 7 days in the SGA3 subgroup (MAE (95% CI) 0·81 (0·72, 0·91) and 0·97 (0·81, 1·15), respectively).

Scatter plots of observed GA versus estimated GA for all three models in the Ontario, Zambia and Bangladesh heel prick cohorts are presented in Figure 3, which shows that in general, lower (preterm) GAs tend to be overestimated by all three models when applied to both external cohorts, with the overestimation being much more pronounced for Model 1 estimates. This pattern was observed in the Ontario test cohort only for Model 1.

**Figure 3:**
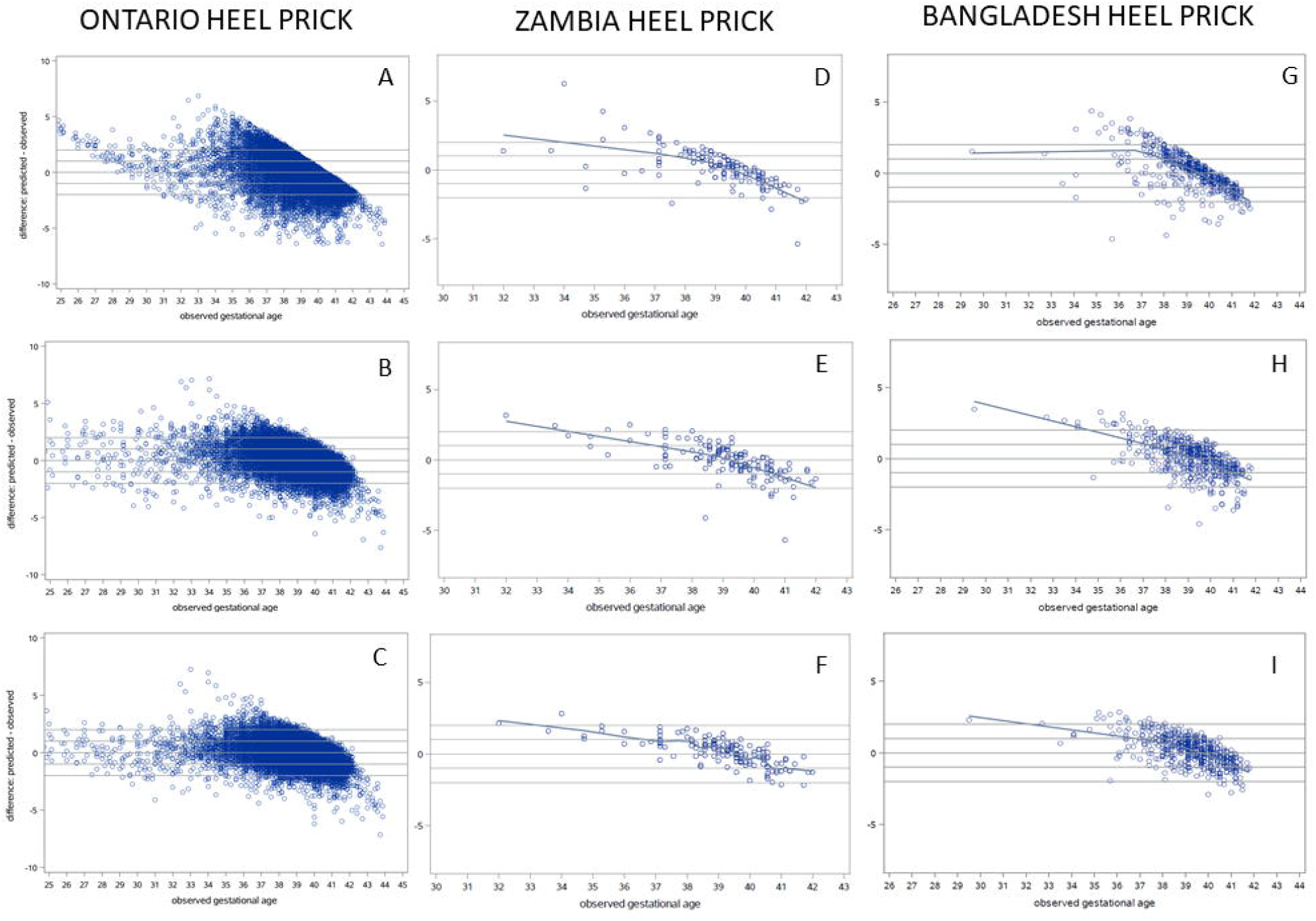
Residual plots of predicted – observed by ultrasound-assigned gestational age. **Heel prick samples from Ontario**: (A) Model 1: Baseline Model, (B) Model 2: Analyte Model, and (C) Model 3: Full Model. **Heel prick samples from Zambia**: (D) Model 1: Baseline Model, (E) Model 2: Analyte Model, and (F) Model 3: Full Model. **Heel prick samples from Bangladesh:** (G) Model 1: Baseline Model, (H) Model 2: Analyte Model, and (I) Model 3: Full Model.

### Model performance using cord blood data from Zambia and Bangladesh

Overall, performance was attenuated when applied to cord blood samples vs. heel prick samples for Models 2 and 3 (the models including analyte covariates), in both the Zambia and Bangladesh cohorts (Table 5 and Figure 2). In the cord blood cohort from Zambia, Models 1 and 3 performed similarly (MAE (95% CI) 1·03 (0·93,1·13) vs. 1·02 (0·90, 1·15)), respectively), and significantly outperformed Model 2 (MAE (95% CI) 1·30 (1·15, 1·47)) (Table 5). Overall, in cord blood samples from Bangladesh, Model 3 significantly outperformed Models 1 and 2 in accurately estimating GA (MAE (95% CI) 0·95 (0·90, 0·99) vs. 1·08 (1·03, 1·13) and 1·06 (1·01, 1·11), respectively).

**Table 5.** Cord blood samples: MAE and proportion estimated within 1 week and 2 weeks of ultrasound-assigned gestational age.

|  | Zambia |  |  |  | Bangladesh |  |  |  |
| --- | --- | --- | --- | --- | --- | --- | --- | --- |
|  | Overall,<br>N=265 | <37 wks,<br>N=22 | SGA10,<br>N=25 | SGA3,<br>N=8 | Overall,<br>N= 1139 | <37 wks,<br>108 | SGA10,<br>N=287 | SGA3,<br>N= 107 |
| **Model 1: Baseline Model** |  |  |  |  |  |  |  |  |
| MAE (95% CI) | **1·03 (0·93, 1·13)** | 2·42 (1·73, 3·12) | 1·52 (1·12, 1·96) | 2·27 (1·40, 3·17) | **1·08 (1·03, 1·13)** | 2·21 (1·98, 2·43) | 1·16 (1·05, 1·27) | 1·86 (1·68, 2·06) |
| % +/-1 wk (95% CI) | **60·0 (54·3, 66·0)** | 36·3 (15·8, 57·1) | 40·0 (20·0, 61·1) | 12·5 (0·0, 40·0) | **56·5 (53·6, 59·2)** | 23·2 (15·5, 31·5) | 54·6 (49·0, 60·3) | 21·5 (13·9, 30·0) |
| % +/- 2 wks (95% CI) | **87·9 (83·8, 91·7)** | 40·8 (20·0, 61·9) | 67·9 (47·8, 86·0) | 37·2 (0·0, 75·0) | **86·1 (84·2, 88·2)** | 41·8 (32·7, 50·9) | 84·6 (80·5, 88·7) | 64·5 (55·2, 73·3) |
| **Model 2: Analyte Model** |  |  |  |  |  |  |  |  |
| MAE (95% CI) | **1·30 (1·15, 1·47)** | 3·96 (2·79, 5·34) | 1·76 (1·14, 2·60) | 2·66 (1·33, 5·03) | **1·06 (1·01, 1·11)** | 2·32 (2·07, 2·58) | 1·11 (1·00, 1·22) | 1·17 (0·97, 1·41) |
| n within 1 wk (95% CI) | **50·2 (43·8, 56·6)** | 10·8 (0·0, 25·0) | 32·5 (13·8, 52·4) | 12·3 (0·0, 40·0) | **56·8 (53·8, 59·8)** | 13·0 (6·9, 19·8) | 51·0 (45·3, 56·7) | 48·7 (38·5, 58·7) |
| n within 2 wks (95% CI) | **81·6 (76·2, 86·4)** | 28·3 (8·5, 50·0) | 68·8 (48·2, 87·5) | 49·8 (11·1, 85·7) | **86·5 (84·5, 88·5)** | 38·9 (29·9, 48·4) | 85·3 (81·1, 89·2) | 85·1 (78·1, 91·5) |
| **Model 3: Full Model** |  |  |  |  |  |  |  |  |
| MAE (95% CI) | **1·02 (0·90, 1·15)** | 3·01 (2·15, 3·99) | 1·15 (0·67, 1·82) | 1·41 (0·25, 3·34) | **0·95 (0·90, 0·99)** | 1·92 (1·74, 2·11) | 0·92 (0·84, 1·00) | 1·10 (0·94, 1·27) |
| % +/-1 wk (95% CI) | **60·7 (54·7, 66·8)** | 13·8 (0·0, 31·3) | 54·6 (33·3, 73·9) | 62·9 (25·0, 100·0) | **61·0 (58·1, 63·8)** | 16·8 (9·8, 23·6) | 60·8 (54·9, 66·4) | 51·4 (41·8, 60·8) |
| % +/-2 wks (95% CI) | **90·1 (86·0, 93·6)** | 38·3 (17·5, 60·9) | 88·0 (71·9, 100·0) | 87·5 (60·0, 100·0) | **91·2 (89·6, 92·8)** | 56·3 (46·7, 65·7) | 92·4 (89·3, 95·3) | 85·1 (78·0, 91·4) |

Data are presented as the mean and 2.5<sup>th</sup> and 97.5<sup>th</sup> bootstrap percentiles for MAE, and the percentage of model estimates within 1 and 2 weeks of ultrasound GA for 2000 bootstrap.

When applied to cord blood samples from preterm infants in Zambia (n=22), all models generally performed poorly. The best performing model in this group was Model 1, with an MAE (95% CI) of 2·42 (1·73, 3·12) and estimated GA within 1 and 2 weeks in 36·3% and 40·8% of samples respectively. Models performed slightly better in estimating preterm GA in cord blood samples from Bangladesh (n=108). Model 3 had the best performance, with a MAE (95% CI) of 1·92 (0·90, 0·99) and was able to correctly estimate GA to within 1 and 2 weeks in 16·8% and 56·3% of preterm infants respectively.

Our validation cohort from Zambia contained a slightly larger number of SGA cord blood samples (25 SGA10 and 8 SGA3 samples) compared to heel prick samples (11 SGA10 and 3 SGA3). Model 3 had the best performance in this subgroup, performing similarly in SGA10 and SGA3 infants and correctly estimating GA to within 2 weeks in 88·0 % (95% CI =71·9,100·0) and 87·5% (95% CI =60·0,100·0) of cord samples, respectively. When applied to cord samples from SGA infants in Bangladesh, Model 3 also had the best performance, and correctly estimated GA to within 2 weeks in 92·4 % (95% CI =89·3,95·3) and 85·1% (95% CI =78·0, 91·4) of cord samples from SGA10 and SGA3 infants, respectively. Figure 4 presents scatter plots for observed versus model-estimated GA for all three models in the Zambia and Bangladesh cord blood cohorts.

**Figure 4:**
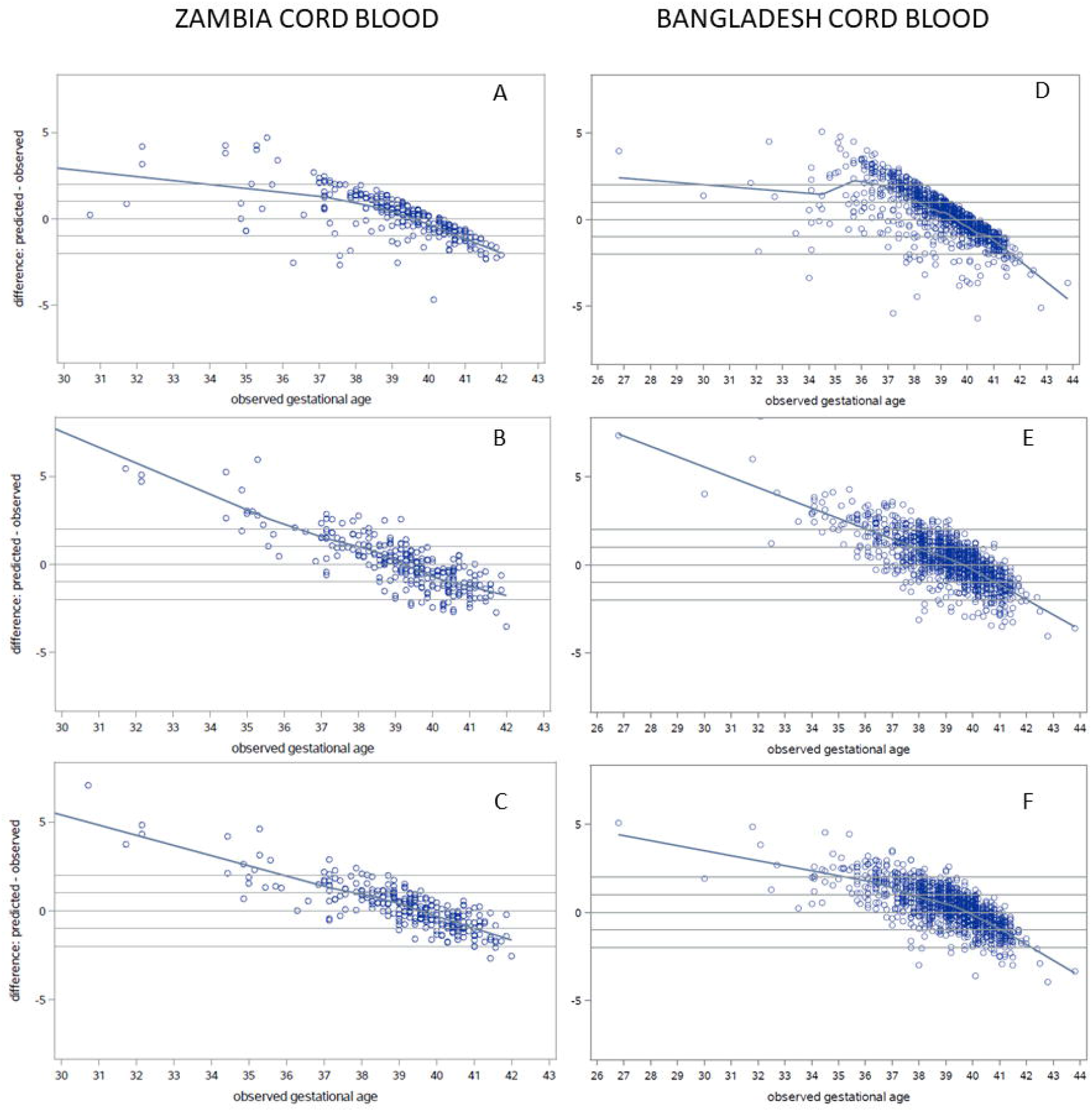
Residual plots of predicted – observed by ultrasound-assigned gestational age. **Cord blood samples from Zambia:** (A) Model 1: Baseline Model, (B) Model 2: Analyte Model, and (C) Model 3: Full Model. **Cord blood samples from Bangladesh:** (D) Model 1: Baseline Model, (E) Model 2: Analyte Model, and (F) Model 3: Full Model.

### Estimated Preterm Birth Rate

In the Ontario cohort, the preterm birth proportion was 5·6% (5·4%, 5·8%) in the testing subset and overall based on ultrasound-determined GA. Model 3, the full model which demonstrated the highest accuracy, estimated the preterm birth proportion to be 5·7% (5·4%, 6·0%) in the Ontario testing subset. In the Zambia heel prick cohort, the observed preterm birth proportion was 7·7% (3·5%, 12·7%) and Model 3 estimated the preterm birth proportion as 5·8% (2·1%, 8·6%). In the Zambia cord blood cohort, the preterm birth proportion was 8·3% (5·1%, 11·7%) and Model 3 estimated the preterm birth proportion to be 4·4% (1·9%, 7·2%). In the Bangladesh heel prick cohort, the observed preterm birth proportion was 6·7% (4·6%, 9·0%) and Model 3 estimated the proportion as 3·8% (2·3%, 5·4%). Likewise, in the Bangladesh cord blood cohort the observed preterm birth proportion was 9·5% (7·9%, 11·2%) and Model 3 yielded an estimated preterm birth proportion of 4·1% (3·0%, 5·3%).

## Discussion

In this study, we demonstrated that our ML algorithms for postnatal GA estimation developed from heel prick blood sample data in Ontario, Canada, can be successfully applied in low and middle income countries in Sub-Saharan Africa and South Asia, having some of the highest preterm birth rates globally ^10,11^. When applied to heel prick samples from Lusaka, Zambia and Matlab, Bangladesh, our GA estimates from Model 3 (our best-performing model) were within an average of 6 days of ultrasound-based GA. All models produced the most accurate estimates in full term infants (37 to 39 completed weeks GA), however Model 3 provided clinically important improvements (by a week or more in some instances) in accuracy over Model 1. Although Model 1’s overall performance was satisfactory, its precision was poor in preterm infants, particularly in growth restricted infants (SGA3 and SGA10) across the spectrum of GA. This is not surprising since Model 1 relies only on sex, birth weight and multiple versus singleton birth to estimate GA. In growth restricted infants especially, birth weight is a misleading measure of GA. Model 3 demonstrated improved accuracy compared to Model 1 in SGA infants across the Ontario, Zambia and Bangladesh cohorts, in most cases with minimal to no overlap in 95% confidence intervals. In Ontario infants, point estimates for Model 3 MAE in SGA10/SGA3 infants were 1·13/1·48 respectively, compared to 2·70/3·85 for Model 1. In Zambia, MAE for SGA10/SGA3 infants was 0·87/1·29 compared to 1·79/3·54 for Model 1. In Bangladesh the MAE for SGA10/SGA3 infants was 0·81/0·97 for Model 3 compared to 1·14/1·73 for Model 1.

Not surprisingly, model performance was sharply diminished when applied to cord blood samples from infants in both external cohorts, however, despite being developed using heel prick samples, Models 2 and 3 still demonstrated some promise in terms of clinical utility in estimating GA using cord blood samples. Because sizable subgroups of the Zambia and Bangladesh cohorts had both a heel prick and cord blood sample collected from the same infants, we were able to investigate the agreement between analyte measurements in the two sample types. We identified analytes with high and low concordance based on Spearman correlation, and derived restricted versions of Model 3 including only analytes meeting a minimum concordance threshold (Online supplement). These restricted models increased the precision of cord blood GA estimates in preterm infants, but not overall (Online supplement).

In our Results, we presented scatter plots of observed versus estimated GA (Figures 3 and 4) that showed a systematic tendency for GA to be overestimated in increasingly preterm infants. This was observed for all three models in heel prick and cord blood samples from Zambia and Bangladesh. The same phenomenon was not observed in the Ontario internal validation results. There are a few possible explanations for this apparent systemic bias which can be framed as a model calibration problem. For example, our reference models employed shrinkage penalization during model fitting (via ELASTICNET regression) to control over-fitting, which may have led to over-penalized final regression model coefficients, leading to the opposite problem of under-fitting. Another potential contributor is the standardization process during data preparation. The same standardization was employed in all three cohorts, but local means and standard deviations were used specific to each cohort. Local standardization was a critical step, and all models had much poorer performance when this step was omitted, however large differences in the dispersion of model predictors across populations, driven by local factors (such as socioeconomic conditions, climate, and underlying differences in birth weight and GA distributions) were likely much larger contributors to differences in covariate distributions. This could have contributed to clinically important variation being muted in the external cohorts. An important implication of this model calibration issue is that preterm (<37 weeks GA) birth estimates based on our models are too low. This is in large part due to “edge effects” which are an important limitation of dichotomizing a naturally continuous variable (for example a GA estimate of 36·9 weeks would be classified preterm while one of 37·1 would be classified as term, despite the estimates being only a day or two apart). Model calibration in new external settings is an important consideration and a focus of ongoing investigation by our research team.

Direct comparisons of model performance across populations and between our current ML GA estimation models and previously reported conventional regression models, is challenging due to different distributions of GAs and birth weights, and other infant- and setting-specific factors. We have not shown a direct comparison of our current models to previous models in this manuscript, however in comparing internal validation results among old and new models, we noted improvements in the accuracy of GA estimates overall, and in preterm/SGA infants. These findings were largely consistent across the internal and external validation cohorts with some variation. Therefore our interpretation is that benefits of our ML approach were incremental, suggesting that our previous models were robustly developed.

Our study had several important strengths. These include our strategy of both internally validating our models in Ontario, as well as our engagement with international collaborators to externally validate in mother-infant cohorts from low and middle income countries. Samples were collected from infants with GA confirmed by early pregnancy ultrasound, and analyzed centrally (in the same lab where samples were analyzed for the Ontario cohort). The ZAPPS and PreSSMat cohorts, in which our study was conducted, ensured that enrollment was open to representative populations of women and newborns in both Lusaka, Zambia and Matlab, Bangladesh. Other strengths include the high quality of samples received and our collection of paired heel and cord blood samples which allowed the comparison of model performance metrics between sample types, as well analyte level comparisons in paired heel prick and cord blood samples from a large subgroup of infants. The superior accuracy of Model 3 (our best performing metabolomic model) in estimating GA in SGA and preterm infants compared to Model 1 (that only relies on clinical variables) is an important strength, given the limitations reported for commonly-used tools in the postnatal period (i.e., Ballard or Dubowitz scores) in these vulnerable infants ^6,7,24^. The primary limitation of this study is the limited number of preterm infants available in our external validation cohorts, especially very and extremely preterm infants (28-32 and <28 weeks gestation respectively). There was a higher likelihood that consent would be provided for the collection of cord blood than for heel prick samples in increasingly preterm infants. This in itself is an important finding as it highlights the importance of further developing estimation models that work well in cord blood samples. In the Bangladesh cohort, parents expressed reluctance to subject their premature newborns to blood sample collection (via heel prick). Although this was not systematically surveyed at the Zambia site, our study nurses reported a similar hesitancy among Zambian parents. Consequently, the relatively small number of heel prick samples collected from very preterm infants limited our ability to interpret model performance in these subgroups. Our nurses also reported a stigma surrounding the heel prick blood sample collection being perceived as associated with early infant diagnosis of HIV. Although the study team focused educational efforts on dispelling this perception, it had a persistent effect on our ability to recruit the targeted number of participants.

Accurate ascertainment of preterm birth rates across LMIC is imperative in order to evaluate the impact of policies and programs aimed at improving pregnancy and neonatal outcomes. In North America, statistical models using data from biochemical analysis of newborn dried blood spots, including those previously developed by our team, have been shown to provide accurate estimates of GA, with some limitations in preterm and growth restricted infants ^12–17^. In this study we have presented internal and external validation results of our most current ML algorithms employing ELASTIC NET regression for GA estimation in both high and low income settings, providing incremental improvements in performance compared to previously developed models. Large-scale implementation of this approach, and population-level collection and analysis of newborn samples, offers a new opportunity to provide surveillance of the burden of preterm birth in jurisdictions where data are currently lacking.

## Data Availability

A de-identified dataset can be provided upon reasonable, written request to the corresponding author and development of appropriate Data Sharing Agreements.

## Author Contributions

KW and SH were responsible for the conception of the study; SH, RD and WC were responsible for data analysis; SH, RD, MSQM, BO, LAW, BKP, JL, MH, and PC contributed to data interpretation; KD, ML, MH, KJR, JTP, HM, BV, PM, JP, AKAC, AR, PC and JSAS were responsible for data acquisition; SH, RD, MSQM and BO wrote the first draft of the manuscript; ABB, LAW, WC, JL, BKP, KD, ML, KJR, PC, JSAS and KW provided critical revisions for important intellectual content. All authors have approved the final version of the manuscript.

